# Deep convolutional approaches for the analysis of Covid-19 using chest X-Ray images from portable devices

**DOI:** 10.1101/2020.06.18.20134593

**Authors:** Joaquim de Moura, Lucía Ramos, Plácido L. Vidal, Milena Cruz, Laura Abelairas, Eva Castro, Jorge Novo, Marcos Ortega

## Abstract

The recent human coronavirus disease (COVID-19) caused by severe acute respiratory syndrome coronavirus 2 (SARS-CoV-2) has been declared as a global pandemic on 11 March 2020 by the World Health Organization. Given the effects of COVID-19 in pulmonary tissues, chest radiography imaging plays an important role for the screening, early detection and monitoring of the suspected individuals. Hence, as the pandemic of COVID-19 progresses, there will be a greater reliance on the use of portable equipment for the acquisition of chest X-Ray images due to its accessibility, widespread availability and benefits regarding to infection control issues, minimizing the risk of cross contamination. This work presents novel fully automatic approaches specifically tailored for the classification of chest X-Ray images acquired by portable equipment into 3 different clinical categories: normal, pathological and COVID-19. For this purpose, two complementary deep learning approaches based on a densely convolutional network architecture are herein presented. The joint response of both approaches allows to enhance the differentiation between patients infected with COVID-19, patients with other diseases that manifest characteristics similar to COVID-19 and normal cases. The proposed approaches were validated over a dataset provided by the Radiology Service of the Complexo Hospitalario Universitario A Coruña (CHUAC) specifically retrieved for this research. Despite the poor quality of chest X-Ray images that is inherent to the nature of the portable equipment, the proposed approaches provided satisfactory results, allowing a reliable analysis of portable radiographs, to support the clinical decision-making process.

## Introduction

Coronavirus disease 2019 (COVID-19) is a respiratory infection caused by the novel coronavirus severe acute respiratory syndrome coronavirus 2 (SARS-CoV-2) (1). It initially emerged in Wuhan, Hubei province, in late December 2019, as a cluster of cases of pneumonia with unknown cause (2), being spread rapidly worldwide. Given the alarming levels of spread and severity as the number of cases of Covid-19 outside China increased drastically as well as the number of affected countries, the World Health Organization (WHO) declared the outbreak of COVID-19 as a global pandemic on 11 March 2020, with more than 118,000 con-firmed cases, including 4,291 deaths reported in 114 countries (3). By May the 13th 2020, it has affected more than 213 countries/regions, with 4,170,424 confirmed cases, including 287,399 deaths, reported to the World Health Organization (WHO) (4).

Among the observable symptoms, the infection can manifest fever, coughing, shortness of breath, pneumonia, and acute respiratory distress syndrome, being associated severe cases with intensive care unit admission and high mortality (5). The severity of the disease is related to advanced age and comorbidities such as chronic heart and lung disease (6). Additionally, the virus is highly contagious, spreading efficiently by human to human transmission via droplets or close contact (7), with an *R*0 ≥ 3, that is, on average each infected person is capable of transmitting the virus to 3 other people (6). Therefore, the COVID-19 is considered as a public health emergence, and according to the key areas described by the WHO, the early diagnosis is a priority in order to break the chain of transmission and, therefore, control the spread of the virus (4).

During the last decades, chest radiography imaging has been a diagnostic tool commonly used in the clinical practice to examine abnormalities in the cardiothoracic region, in order to detect and monitor the evolution of several pulmonary diseases, such as emphysema, chronic bronchitis, pulmonary fibrosis, lung cancer or pneumonia, among others (8–10). Regarding the COVID-19 disease, several manifestations and patterns of lung abnormality were identified in chest X-Ray images such as bilateral abnormalities, interstitial abnormalities, lung consolidation and ground glass opacities (11). Therefore, the analysis of chest X-Ray images of the suspected individuals presents a remarkable potential for screening processes and early diagnosis of COVID-19 disease (12), even more when it is known that Polymerase Chain Reaction (PCR) studies, despite their effectiveness, have a percentage of false negatives, being therefore the radiological analysis another complementary tool to support and confirm the diagnosis of these patients (13). In this context, the specialist’s experience is essential for the identification and characterization of biomarkers associated with COVID-19, as well as the differentiation from other clinical findings related to other respiratory diseases with similar characteristics, as pneumonia. However, the subjective appreciation of the clinical biomarkers and the variety of X-Ray images entail an inter and intra expert variability that affects the repeatability of the manual evaluation of chest X-Ray images. Besides the subjectivity, the manual characterization is a tedious and time consuming task. Therefore, a fully automatic system for the analysis of chest X-Ray images identifying normal, pathological or specific COVID-19 cases would reduce the workload of the clinical staff allowing a reliable and repeatable evaluation to support the clinical decision-making process.

Over the recent years, deep learning techniques have demonstrated their potential, effectiveness and versatility, positioning themselves as benchmarks in solving a large number of problems in multiple fields, highlighting computer vision. Given the relevance of the topic, during the last months, several authors proposed computational approaches for the COVID-19 classification on chest X-Ray images using deep learning techniques. As reference, Narin *et al*. (14) evaluates three different approaches using convolutional neural networks based on transfer learning for the detection of pneumonia associated with COVID-19 in chest X-Ray images. Apostolopoulos *et al*. (15) explore the possibility of extracting representative COVID-19 biomarkers from chest X-Ray images by means of deep learning strategies. In the work of Wang *et al*. (16), the authors present COVID-Net, a deep convolutional neural network tailored designed for detecting COVID-19 cases from chest X-Ray images. Hammoudi *et al*. (17) proposed a deep learning method applied to chest X-Ray images to identify health and pneumonia cases, differentiating between bacterial or viral origin, assuming that, during the epidemic season, if the origin of pneumonia is viral, there is a high probability of being a positive COVID-19 case. De Moura *et al*. (18) presented several fully automatic deep convolutional approaches for the classification and study of COVID-19, common pneumonia and healthy chest X-Ray images. In the work of Zhang *et al*. (19), the authors proposed a deep anomaly detection model for COVID-19 screening on chest X-Ray images. Ozturk *et al*. (20) proposed a deep learning model that provides heatmaps highlighting important areas in chest X-Ray images to help the specialists to locate the affected regions for the early detection of COVID-19 disease.

Although the methods that were proposed in the literature provide satisfactory results, these approaches were trained using datasets composed by chest X-Ray images acquired by fixed X-Ray equipment, presenting high quality in the produced images. However, in the context of the global pandemic caused by COVID-19, a properly decontamination of the equipment used for the acquisition of the chest X-Ray images as well as an effective control during the patient transport to the X-ray room are critical factors in controlling the transmission of the virus (21). The use of portable equipment for the acquisition of chest X-Ray images allows a more effective separation of the circuit of the suspected patients, avoiding to share infected patients with those without suspected infection by COVID-19, either in the radiology room itself, or in common areas as waiting rooms or hallways. In this sense, the American College of Radiology (ACR) recommends the use of portable chest X-Ray acquisition equipment in order to minimize the risk of cross infection given the ease of decontamination of the surface of portable devices and the capability of acquiring the chest X-Ray images without the necessity of taking the patient to the X-Ray room (22). Therefore, portable radiological studies have been the first and main radiological analysis procedure in order to detect possible findings that support the diagnosis of suspected patients, as well as determine its severity, minimizing the risk of virus transmission. Additionally, portable devices can also be very effective for the triaging of patients with early symptoms but instructed to stay at home if there are no severe signs, thus avoiding the risk of virus spreading and the collapse of hospital centers. Furthermore, as the COVID-19 pandemic progresses, there will be a greater reliance on portable devices given their accessibility, widespread availability as well as the reduced decontamination issues required for an adequate use (21).

Given the great relevance and utility of the portable equipment to face the public health emergency caused by the COVID-19 pandemic, in this work, we propose a comprehensive methodology and derived accurate approaches for the analysis of COVID-19 in chest X-Ray images acquired by portable devices. Considering that several respiratory affections can present clinical findings similar to COVID-19, hindering the differentiation between patients that have contracted COVID-19 of those that are affected by other respiratory diseases, the proposed methodology integrates two complementary approaches based on deep learning techniques. The first of these approaches is aimed at classifying between normal patients and those pathological cases that present any indication that could be related with COVID-19, either specifically related to the presence of COVID-19 or other respiratory conditions with similar symptoms. Additionally, the second approach is specifically trained for the differentiation of the patients with COVID-19 with respect to patients not infected with COVID-19, whether normal cases or with respiratory conditions other than COVID-19, but with similar symptoms, the most complex differentiation scenario.

Thus, the joint response of both complementary approaches allows reinforcing the differentiation between COVID-19, pathological patients with other similar respiratory conditions as well as other normal cases.

The proposed methodology and designed approaches were validated over a dataset provided by the Radiology Service of the Complexo Hospitalario Universitario A Coruña (CHUAC) that provided the assistance work of the radiodiagnostic and computer services in order to search for adapted solutions for the improvement in patient care.

Summarizing, the main contributions of the article include: (i) a new computational approach for the classification of chest X-Ray images acquired by portable devices into 3 categories: normal, pathological and COVID-19. The methodology integrates two complementary deep learning approaches in order to enhance the differentiation of COVID-19 cases both from patients with pulmonary conditions that manifest clinical findings similar to COVID-19 and from normal patients without any similar abnormality in the lung tissues. We would like to highlight that the considered pathological cases are those from pulmonary diseases with similar findings as COVID-19, whereas the normal subjects may not specifically healthy. In that case, they belong to healthy patients as well as other pathological cases with different appearance with characteristics to COVID-19. In that line, we selected in this work the most complex scenario of differentiation to analyze the COVID-19 disease; (ii) to date, this proposal represents the only study specifically designed for the analysis of COVID-19 in chest X-Ray images acquired by means of portable devices, with a significant penalization in the quality of the captured images; (iii) the methodology has been trained and validated using a dataset specifically designed for this research; (iv) these fully automatic approaches provided accurate results even though portable devices acquire images with poor quality conditions, allowing a reliable analysis to support the clinical decision-making process in the context of this dramatic global pandemic.

The manuscript is structured as follows: Section presents the dataset and describes the different deep learning approaches that were proposed in this research. Next, Section E exposes all the conducted experiments whereas Section G discusses the obtained results. Finally, Section G presents the conclusions that are extracted from this research as well as possible future work is the analysis of this relevant disease.

## Materials and Methods

### A. Used image dataset

The used dataset was provided by the Radiology Service of the Complexo Hospitalario Universitario A Coruña (CHUAC). These images have been acquired by means of portable equipment, specifically, Agfa dr100E GE and Optima Rx200 portable devices. These portable devices were located in areas specifically intended for COVID-19 in order to examine patients referred after undergoing triage in the emergency systems, and monitor these patients in the different hospital plants. The use of portable equipment allows an effective separation between infected patients and patients without suspected COVID-19, both in the radiology room itself and in waiting rooms or hallways. For the acquisition procedure, the patient lies in supine position and a single anterior-posterior projection is recorded. For this purpose, the X-Ray tube is connected to a flexible arm that is extended over the patient to be exposed to a small dose of ionizing radiation while an X-Ray film holder or an image recording plate is placed under the patient to capture images of the interior of the chest.

The dataset consists of 1,616 chest X-Ray images divided into 728 normal cases corresponding to patients who have neither pleural nor pulmonary pathologies (but other pathologies have not been assessed, such as cardiology or hepatic diseases, for example) at the time of image acquisition, 648 pathological cases corresponding to patients without COVID-19 but diagnosed with other pulmonary diseases that can present characteristics similar to COVID-19, as well as 240 specific COVID-19 cases. Therefore, this dataset provides a very challenging scenario, with a pathological category with diseases that are similar in terms of characteristics to COVID-19 and normal subjects that could present other non related abnormalities. In any case, the dataset presents cases that are very frequent in real clinical practice scenarios. Fig. 1 shows representative examples of chest X-Ray images related to the presented 3 clinical categories: normal, pathological and COVID-19.

**Fig. 1.**
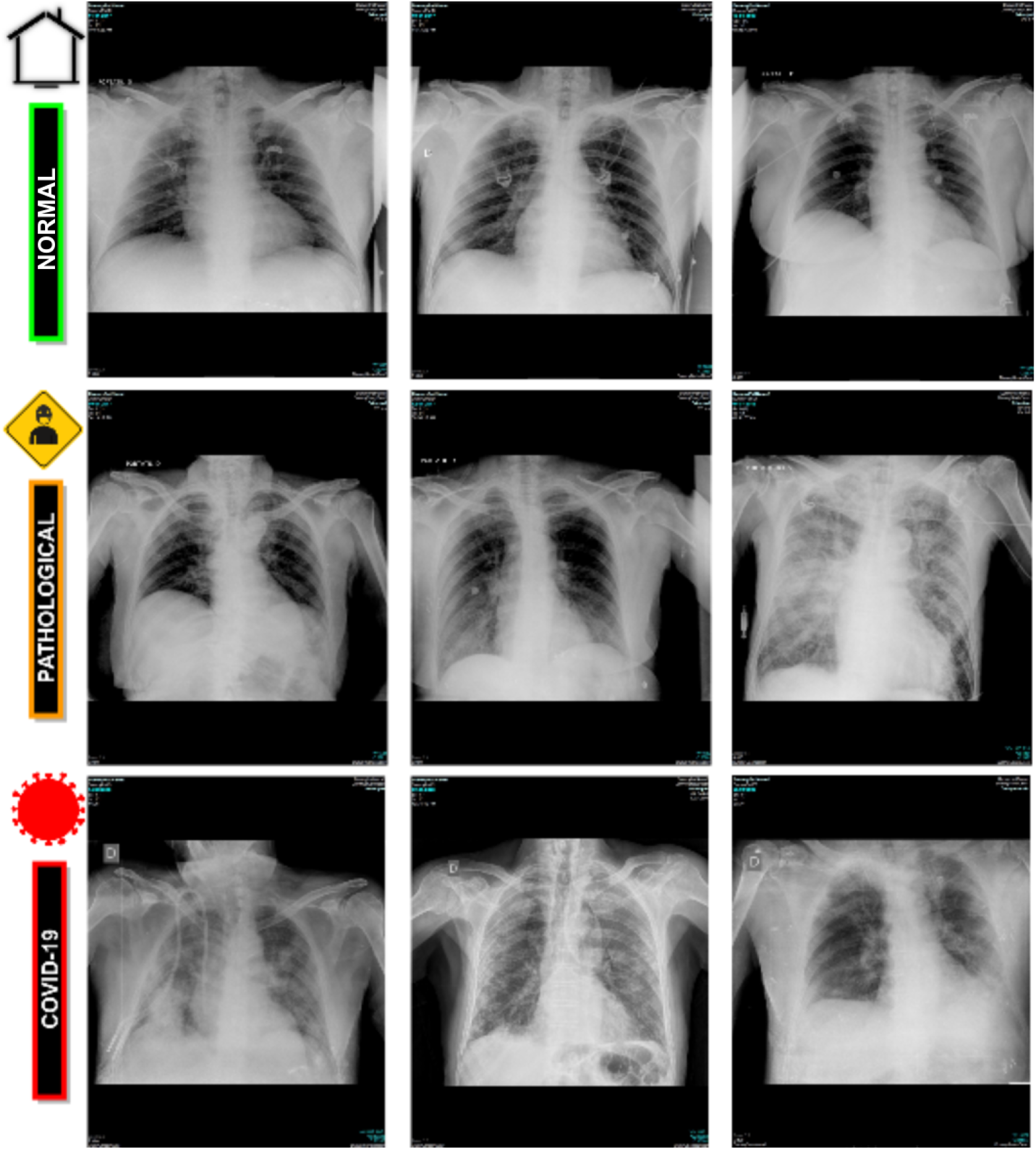
Representative examples of chest X-Ray radiographs. 1^*st*^ row, chest X-Ray radiographs from normal patients. 2^*nd*^ row, chest X-Ray radiographs from pathological patients without COVID-19 but diagnosed with other pleural or pulmonary diseases. 3^*rd*^ row, chest X-Ray radiographs from patients with COVID-19.

### B. Computational approaches for the classification of chest X-Ray images

The impact of COVID-19 on lung tissues can present similarities with other patterns associated to some pulmonary pathologies as common types of pneumonia, especially, during the early stages of both respiratory conditions. In order to enhance the differentiation of COVID-19 cases both from cases that present clinical findings similar to COVID-19 and from patients without respiratory conditions, 3 different clinical categories were designed and considered. Thus, the category *Normal* correspond to patients without pleural or pulmonary pathologies, the category *Pathological* is related to patients diagnosed with pleural or pulmonary pathological conditions other than COVID-19, and the category *COVID-19* is specific for COVID-19 infected patients, as stated in Section A.

A general overview of the proposed computational methodology is depicted in Fig. 2. It receives a chest X-Ray image captured by the portable device as input and provides its classification into the 3 defined clinical categories: *Normal, Pathological* or *COVID-19*. The proposed paradigm is exploited in two complementary approaches based on deep learning techniques for the classification of chest X-Ray images. Both proposed approaches, herein presented, are detailed below:

**Fig. 2.**
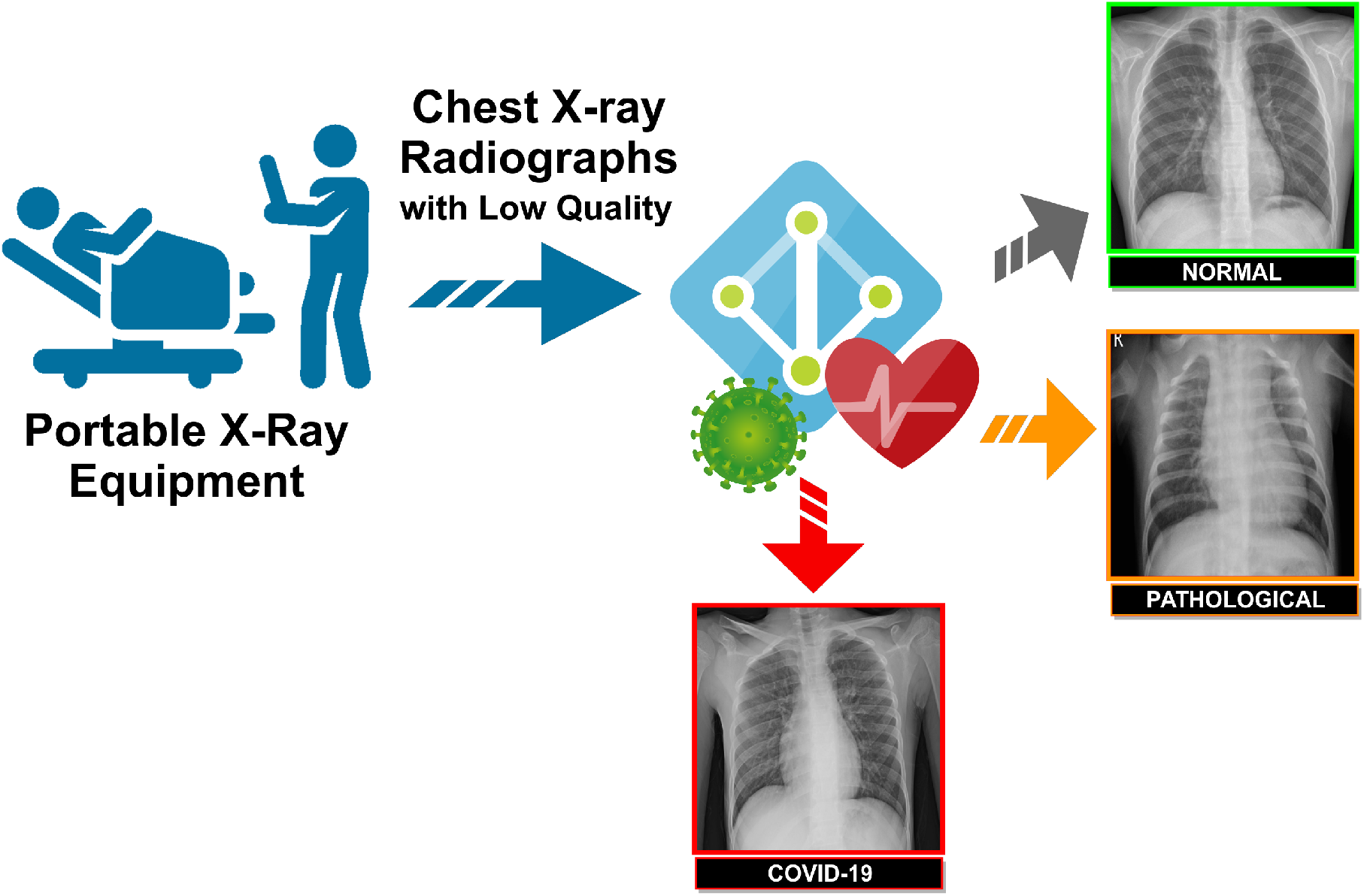
A general overview of the proposed methodology.

#### B.1. Normal vs Pathological/COVID-19 approach

Given the similarities between COVID-19 and other respiratory conditions, the first trained approach is aimed at differentiating between patients with any abnormality in the pulmonary tissues and patients with clinical findings in the lungs, either due to COVID-19 or other respiratory conditions. To this end, chest X-Ray images associated with pathological or COVID-19 cases are grouped under the same class for the training procedure. Therefore, this approach predicts 2 different categories: *Normal* and *Pathological/COVID-19*.

#### B.2. Normal/Pathological vs COVID-19 approach

The second trained approach is specifically intended to identify COVID-19 cases, differentiating them not only from normal cases, but also from those pathological patients affected by other respiratory conditions that manifest similar characteristics, such as pneumonia. For this purpose, chest X-Ray images from normal patients and those from pathological patients with respiratory conditions other than COVID-19 are grouped in the same class, and, therefore, the approach predicts *Normal/Pathological* and *COVID-19* categories.

### C. Network Architecture

Over the recent years, deep learning methods have demonstrated to be an excellent technique in the area of artificial intelligence given its potential and ability to recognise complex patterns from raw input data, learning proper hierarchical representations of the underlying information at different levels. The great potential of these techniques is increasingly used in many relevant fields of medical imaging (23, 24), including computer-aided detection/diagnosis systems and medical image analysis for accurate early detection, diagnosis, treatment and monitoring of many relevant diseases (25, 26).

In this work, we exploit the potential of a densely convolutional network architecture inspired by the DenseNet proposal of Huang *et al*. (27), which was adapted to our issue due to its flexibility and simplicity and its preceding promising results in other classification tasks applied to the diagnosis of pulmonary diseases (28–30). For this research, the original structure of the DenseNet-161 architecture has been adapted, as depicted in Table 1. Specifically, in this architecture, each layer is connected to every other layer in a feedforward fashion within each dense block to ensure maximum information flow between layers. In order to preserve the feed-forward nature, each layer processes feature maps from all the preceding layers as separate inputs and transfers its own feature maps to all subsequent layers. The classification layer has been adapted to support the output defined for each deep learning approach considered by this proposal, that is, the categorization of chest X-Ray images into 2 clinical classes, considering normal or pathological categories in the first approach and Non-COVID-19 or COVID-19 categories, in the second approach.

**Table 1.** An illustration of the DenseNet-161 structure that was adapted for the experiments of this work.

| Layers | Convolution | Pooling | Dense block | Transition layer |  | Dense block | Transition layer |  | Dense block | Transition layer |  | Dense block | Classification layer |  |
| --- | --- | --- | --- | --- | --- | --- | --- | --- | --- | --- | --- | --- | --- | --- |
| Output Size | 112×112 | 56×56 | 56×56 | 56×56 | 28×28 | 28×28 | 28×28 | 14×14 | 14×14 | 14×14 | 7×7 | 7×7 | 1×1 | - |
| Structure | 7×7<br>Convolution<br>Stride 2 | 3×3<br>Maxpool<br>Stride 2 | $\begin{bmatrix} 1 \times 1 \text{ conv} \\ 3 \times 3 \text{ conv} \end{bmatrix} \times 6$ | 1×1<br>Convolution | 2×2<br>Average<br>pool<br>Stride 2 | $\begin{bmatrix} 1 \times 1 \text{ conv} \\ 3 \times 3 \text{ conv} \end{bmatrix} \times 12$ | 1×1<br>Convolution | 2×2<br>Average<br>pool<br>Stride 2 | $\begin{bmatrix} 1 \times 1 \text{ conv} \\ 3 \times 3 \text{ conv} \end{bmatrix} \times 36$ | 1×1<br>Convolution | 2×2<br>Average<br>pool<br>Stride 2 | $\begin{bmatrix} 1 \times 1 \text{ conv} \\ 3 \times 3 \text{ conv} \end{bmatrix} \times 24$ | 7×7<br>Global<br>average<br>pool | Fully-connected<br>softmax<br>2/3 classes |

### D. Training details

The dataset was randomly divided in three mutually exclusive sets, trying to balance the number of chest X-Ray images associated to each category along the sets. In particular, this partition includes 60% of the cases for training, 20% for validation and the remaining 20% for testing.

For the training procedure, the weights from a model pretrained on the ImageNet dataset have been used for the dense network architecture initialization (31). Thus, the pretrained model allows a more efficient learning process and fast convergence since the weights are initially stabilized. Additionally, it significantly reduces the amount of labeled data required for an adequate model training, as is our case.

Each presented deep learning approach is trained end-to-end with a cross-entropy loss function (32) over the output class and the ground truth for the target chest X-Ray image. The optimization during the training stage is performed by means of Stochastic Gradient Descent (SGD) with a constant learning rate of 0.01, a mini-batch size of 4 and a first-order momentum of 0.9. This optimizer, despite its simplicity, is highly efficient for the discriminative learning of classifiers under convex loss functions (33), defined in Equation 1:

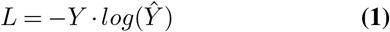

where *Y* represents the ground truth values and Ŷ represents the estimated values for each identified category. A complete training epoch includes a forward pass of all the target frames composing the training set.

In order to assess the generalisation capability of the presented deep learning approaches to extrapolate to unseen chest X-Ray images, 5 repetitions of the whole process were performed, being calculated the mean cross-entropy loss and the mean accuracy to evaluate their global performance.

#### D.1. Data Augmentation

Despite the pretraining, given the dimensionality of the dataset, data augmentation was performed in the experiments, increasing the number of the training samples in order to avoid overfitting and improve the generalization of the network, obtaining more robust and stable trained models (34, 35).

Thus, given the limited amount of COVID-19 subjects, different configurations of affine transformations are applied to the input chest X-Ray images to increase the variability of the training data and improve the performance of the convolutional neural networks. In particular, considering the possible resolutions provided by the common variety of acquisition devices as well as the the symmetrical nature of the human body, several combinations of scaling with horizontal flipping operations were included throughout the training procedure.

### E. Evaluation

The performance of the presented approaches was evaluated by comparing the predictions provided by the networks with the ground truth labels annotated in the dataset. Then, using as reference the True Positives (TP), True Negatives (TN), False Positives (FP) and False Negatives (FN) extracted from this comparison, several performance metrics, commonly used in the literature to evaluate the suitability of computational methods for medical imaging, were considered. Thus, Precision, Recall, F1-score and Accuracy were computed for analyzed approach as follows:

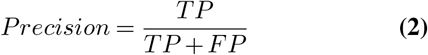

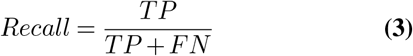

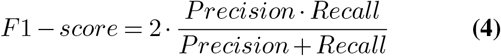

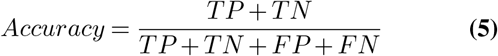

## Experimental Results

In this section, we present the experimental results of the proposed approaches for the classification of COVID-19 in chest X-Ray images. For this purpose, we considered 2 complementary experiments in order to explore the full potential of the available dataset. In this sense, for each proposed experiment, we performed 5 independent repetitions with a different random selection of the sample divisions. In particular, as said, each selection includes 60% of the cases for training, 20% for validation and the remaining 20% for testing. In order to train the adapted DenseNet architecture, we used the cross-entropy loss as training loss function and SGD optimizer with learning rate of 0.01, a mini-batch size of 4 and a first-order momentum of 0.9. In addition, the training process was stopped after 200 epochs due to the lack of significant improvements in both cross-entropy loss and classification accuracy performances.

### F. 1^*st*^ experiment: analyzing the Normal vs Pathological/COVID-19 approach

The first scenario was designed in order to evaluate the performance of the proposed approach to distinguish between chest X-Ray images of subjects with COVID-19 from other similar pulmonary or pleural pathological conditions as well as other non related normal subjects. Accordingly, we designed an experiment with a total of 1,616 chest X-Ray images, being 240 from COVID-19, 648 from patients with other similar lung diseases and 728 from normal subjects. Figs. 3 and 4 show the performance that was achieved using the DenseNet architecture after 5 independent repetitions in the training and validation stages in terms of mean ± standard deviation training accuracy and mean ± standard deviation training loss, respectively.

**Fig. 3.**
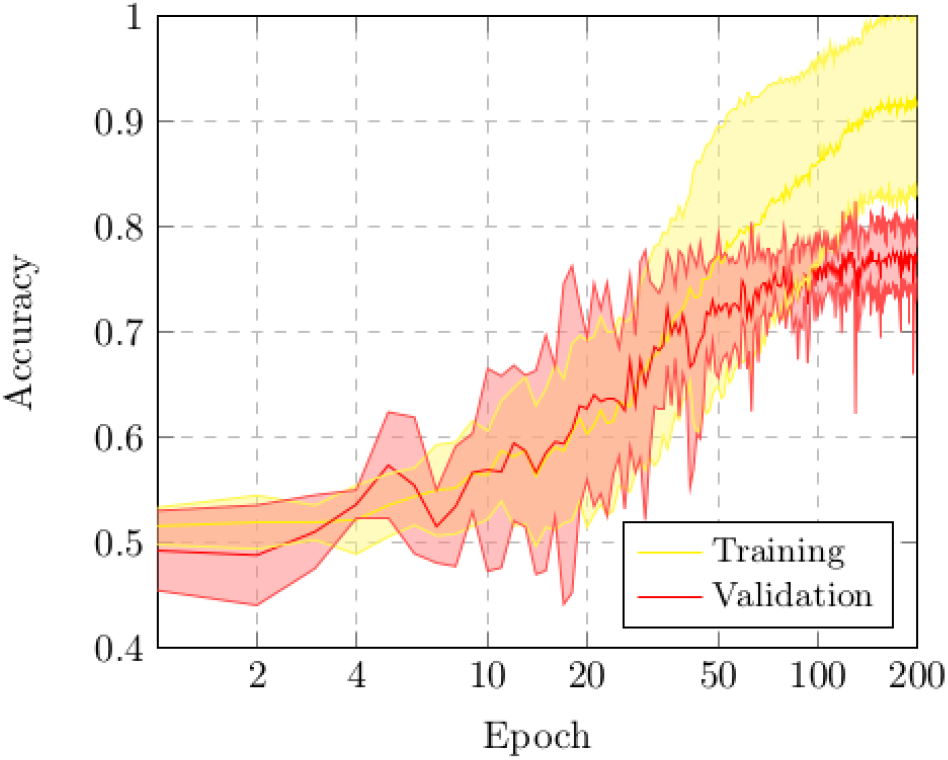
Mean ± standard deviation training and validation accuracy after the 5 independent repetitions of the first experiment. A logarithmic scale has been set to correctly display the values for a better understanding of the results.

**Fig. 4.**
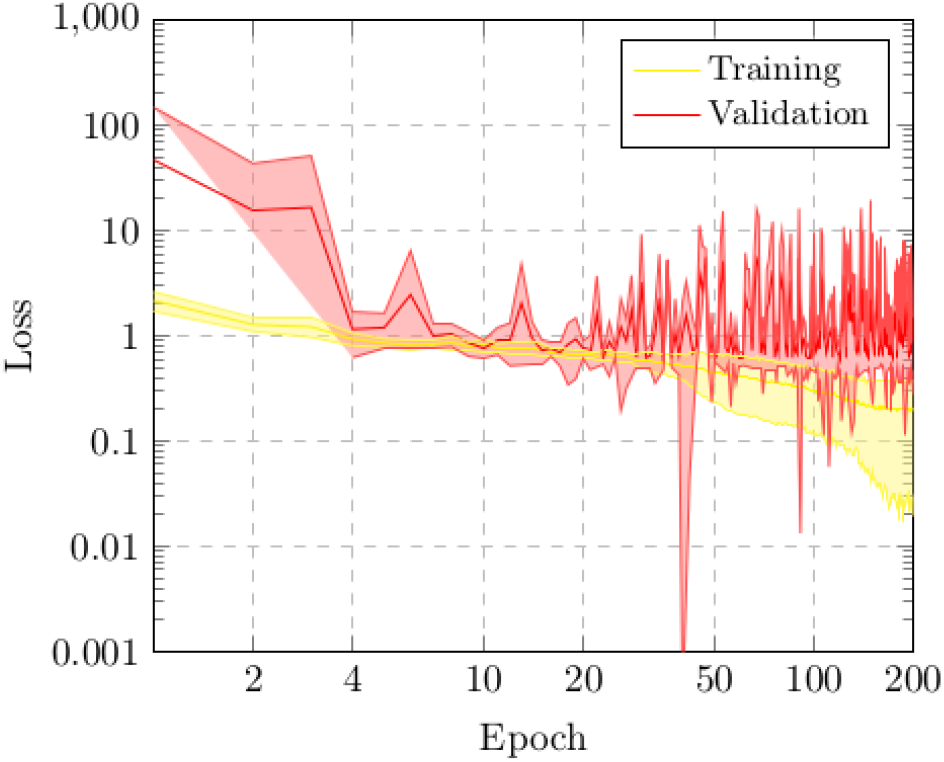
Mean ± standard deviation training and validation loss after the 5 independent repetitions of the first experiment. A logarithmic scale has been set to correctly display the values for a better understanding of the results.

In particular, as we can observe in Fig. 3, the best mean accuracy obtained is 0.9219 ± 0.0810 for the training stage in epoch 194 and the best average accuracy for the validation stage of 0.7783 ± 0.0260 in epoch 144. Additionally, the proposed approach achieved its stability in the cross-entropy function loss both for training and for validation after epoch 150, as we can see in Fig. 4.

Table 2 presents a comparative analysis using precision, recall and F1-score measures. As we can see, the proposed strategy provided satisfactory results, reaching a global accuracy of 79.62%, using all the chest X-Ray images of the analyzed test dataset.

**Table 2.** Precision, recall and F1-score results obtained at the test stage for the classification of chest X-Ray images between Normal vs Pathological/COVID-19 cases.

| Cases | Precision | Recall | F1-score |
| --- | --- | --- | --- |
| **Normal** | 0.74 | 0.82 | 0.78 |
| **Pathological/COVID-19** | 0.85 | 0.78 | 0.81 |

We would like to remark that the pathological combined with the COVID-19 cases present a variety of diseases that presents a significant variability of conditions in the lungs. Also, normal subjects are those from healthy patients but also from other pathological scenarios with characteristics different from COVID-19. Given that, we have to consider the significant variability of characteristics that are represented in both considered cases of this approach. As said, given those circumstances as well as the low quality of the chest X-Ray images captured by portable devices, we consider that these results are satisfactory. Therefore, the results obtained show that this proposal is able to adequately distinguish pulmonary pathological cases from normal patients.

### G. 2^*nd*^ experiment: analyzing the Normal/Pathological vs COVID-19 approach

In this second experiment, we designed a complementary scenario to evaluate the ability of the second analyzed approach to distinguish between cases of patients with COVID-19 from the rest of similar respiratory diseases or even normal patients.

With this in mind, the proposed approach was validated using a dataset composed by 720 chest X-Ray images, being 240 from normal patients whereas other 240 from pulmonary diseases and 240 from COVID-19 patients, respectively. This way, we respect a proportion of 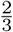 and 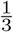 for the Normal/Pathological and COVID-19 classes, trying to keep a minimum of balance between both negative and positive classes of this analyzed approach. In this case, those subjects from pulmonary diseases and normal patients were randomly selected from the total available in the used image dataset.

Figs. 5 and 6 illustrate the performance that was obtained from the proposed deep neural architecture after 5 independent repetitions in both the training and validation stages. In particular, the approach achieved satisfactory results, reaching a best average accuracy of 0.9550 ± 0.0204 for training in epoch 166 and the best average accuracy for validation of 0.8902 ± 0.0158 in epoch 131, as we can see in Fig. 5. In addition, the proposed approach achieved its stability in the loss cross-entropy function both for training and for validation after epoch 100, as we can see in Fig. 6.

**Fig. 5.**
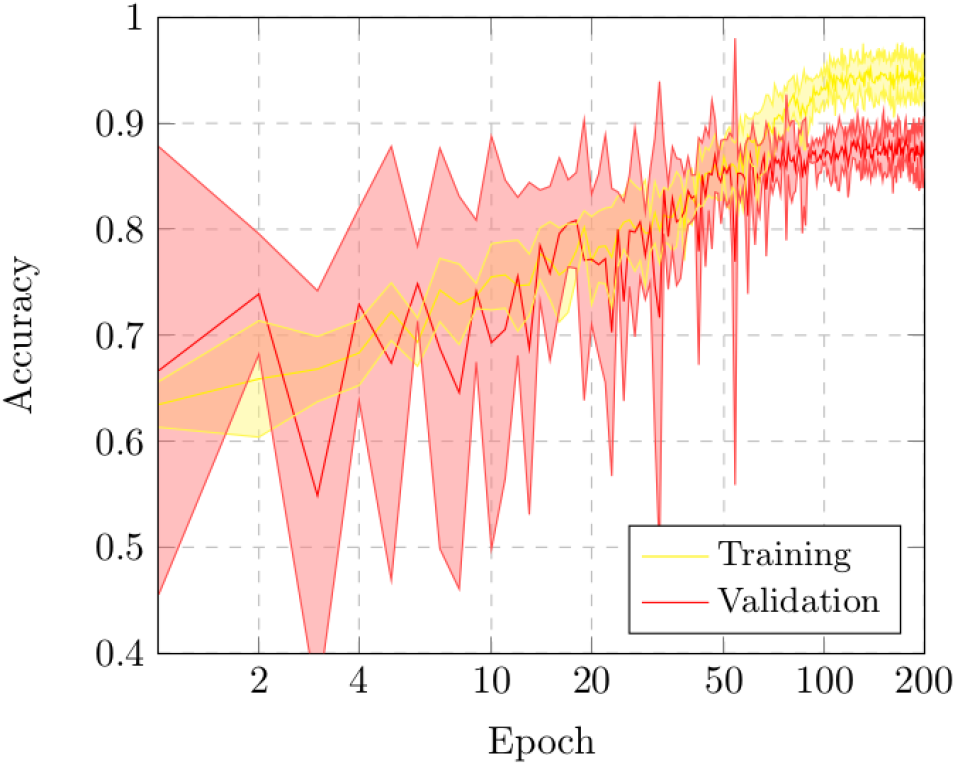
Mean ± standard deviation training and validation accuracy after the 5 independent repetitions of the second experiment. A logarithmic scale has been set to correctly display the values for a better understanding of the results.

**Fig. 6.**
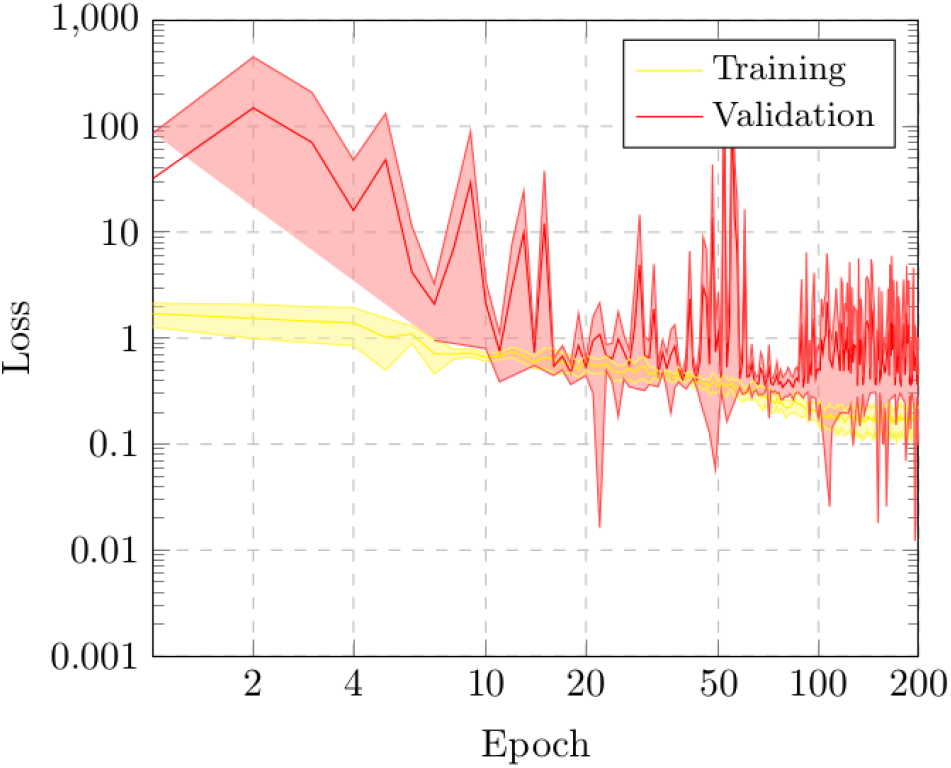
Mean ± standard deviation training and validation loss after the 5 independent repetitions of the second experiment. A logarithmic scale has been set to correctly display the values for a better understanding of the results.

Table 3 summarizes the performance measurements that were obtained by the proposed approach using the test dataset, in terms of precision, recall and F1-score for each category. As we can see, our approach shows a satisfactory performance, providing a global accuracy of 90.27%. In general, the results are in line with the training performance being the proposed approach able to satisfactory classify chest X-Ray images between cases of patients with COVID-19 from other similar pulmonary pathologies or normal subjects. In this case, we have to consider that the separability is more feasible. In particular, the positive class only contains COVID-19 patients, with more restricted characteristics, being the network capable of distinguishing them from the rest of subjects, outperforming the results of the first approach.

**Table 3.** Precision, recall and F1-score results obtained at the test stage for the classification of chest X-Ray images between Normal/Pathological vs COVID-19 cases.

| Cases | Precision | Recall | F1-score |
| --- | --- | --- | --- |
| **Normal/Pathological** | 0.91 | 0.95 | 0.93 |
| **COVID-19** | 0.88 | 0.80 | 0.84 |

## Discussion

Chest X-Ray has become the first-line imaging modality used for the identification of patients with COVID-19 infections at an early stage when clinical symptoms may be nonspecific or sparse. In this context, portable radiography devices are widely used by clinical specialists to evaluate suspected or confirmed cases of COVID-19 disease directly on site, without the necessity of transferring patients potentially infected by COVID-19 elsewhere. Therefore, this clinical procedure helps to save critical time and resources for the therapeutic process as well as reduce the risk of cross-infection.

The use of portable equipment implies a greater challenge for the automatic diagnostic of the COVID-19, given that the acquired images present lower quality, being in the supine position, providing a single anterior-posterior view, unlike radiological analysis carried out in conventional rooms that supplies 2 projections, anterior-posterior and lateral. Despite the poor quality conditions of chest X-Ray images acquired by portable equipment, the obtained results show that the proposed approaches allow a robust and reliable analysis to support the clinical decision-making process in a real public health emergency scenario.

In order to provide a visual interpretation of the proposed approaches, we used the Gradient Class Activation Map (Grad-CAM) (36) algorithm to generate the class activation maps of the trained models, showing us what the neural network sees and what it values when making its prediction. As result, the trained models were shown to be strongly correlated with the clinical findings, as validated by the clinical experts. In Fig. 7, we can see some representative examples of the generated maps based on predictions that illustrate parts of the chest X-Ray image that are strongly activated, as well as a considerable variability of the possible scenarios represented in this research work such as the poor contrast of the radiographs or the great similarity between COVID-19 and others pulmonary diseases that are present in our dataset.

**Fig. 7.**
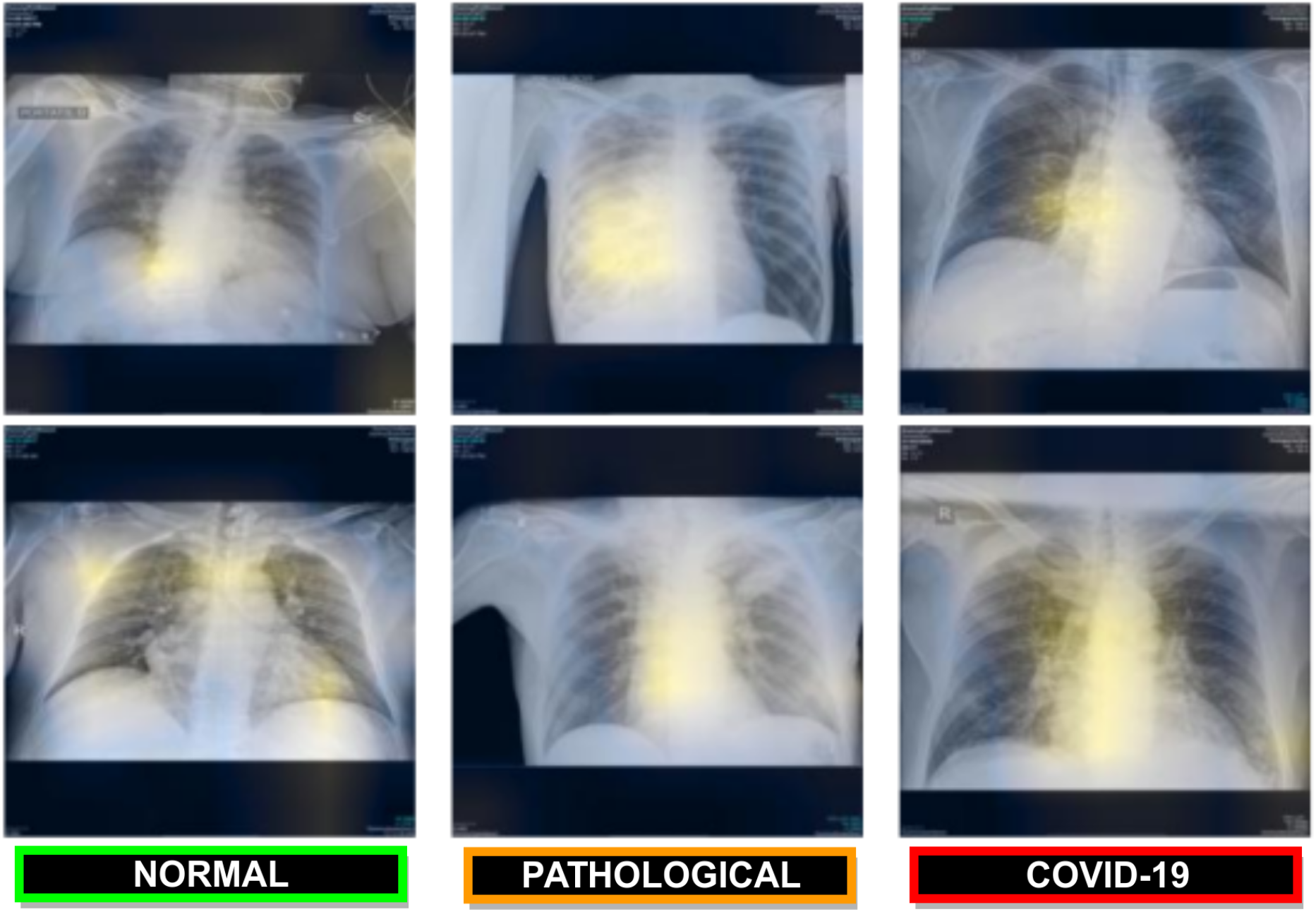
Representative examples of the generated gradient class activation maps based on predictions that illustrate parts of the chest X-Ray image that are strongly activated. 1^*st*^ row, using the Normal vs Pathological/COVID-19 approach. 2^*nd*^ row, using the Normal/Pathological vs COVID-19 approach.

To the best of our knowledge, these are the first fully automatic approaches that was designed specifically for the classification of COVID-19 using chest X-Ray images acquired only by portable equipment. Therefore, we cannot make a direct comparison with other state-of-the-art methods.

From the obtained results, we can indicate that both tested approaches offered satisfactory results. In both scenarios, we have to consider the complexity of analyzing chest X-Ray images capture by portable devices, which present a lower quality in comparison with other typically used images in many pulmonary computational proposals using X-Ray images. This poor quality is combined with the complex tested scenarios of the dataset, considering images from pulmonary conditions that may be confused with the characteristics of COVID-19 as well as normal subjects that include healthy subjects but also may include other pathological conditions that are not related with the characteristics of COVID-19. As said, the designed dataset presents cases that are very frequent in real clinical practice scenarios.

Considering these complex scenarios of differentiation and the poor quality of the analyzed images, the capabilities of the adapted convolutional neural network offered an adequate performance, distinguishing specifically the COVID-19 cases from the rest, but also differentiating both COVID-19 and pulmonary pathological cases from other normal cases, complex scenario with a large variability in both negative and positive classes.

## Conclusions

Coronavirus disease (COVID-19), caused by the newly discovered severe acute respiratory syndrome coronavirus 2 (SARS-CoV-2) is posing high pressure on healthcare services worldwide given its drastic spreading. Therefore, the screening, early detection and monitoring of the suspected individuals are crucial to control the virus transmission. Given the impact of COVID-19 in the pulmonary tissues, chest radiography plays a main role for the examination of the manifestations and patterns of lung abnormality associated with the disease in order to support the diagnosis and determine its severity. Furthermore, in the context of this global pandemic, the use of portable equipment for the acquisition of chest X-Ray images represents the first and main radiological way of analysis due to the more effective separation of the infected patients, allowing to minimize the infection control issues and, therefore, the risk of cross contamination.

Thus, this work proposes a novel fully automatic methodology for the analysis of chest X-Ray images acquired by means of portable devices. Given the similarities between COVID-19 and other pulmonary diseases that also affect the lungs, the proposed methodology is composed of two complementary deep learning approaches in order to better analyze and differentiate different patients infected with COVID-19, patients with other pathologies with similar characteristics to COVID-19 and normal patients. Thus, on one hand, the first approach allows to classify between normal and pathological patients, being pathological both patients with COVID-19 and patients with other similar respiratory conditions. On the other hand, the second approach is aimed at differentiating between patients with COVID-19 and patients that may be normal or present respiratory conditions other than COVID-19. Thus, the joint response of both approaches allow to classify and better analyze the considered COVID-19, pathological and normal cases.

The proposed methodology and both tested approaches were validated using a dataset provided by the Radiology Service of the Complexo Hospitalario Universitario A Coruña (CHUAC). To the best of our knowledge, it is the first proposal specifically tailored for the classification of chest X-Ray images acquired by portable equipment. Despite the poor quality conditions of these chest X-Ray images acquired by portable equipment, the obtained results show that the propose methodology and both tested approaches allow a robust and reliable analysis to support the clinical decision making process in this context of public health emergency.

## Data Availability

The data is not yet available.

## Acknowledgement

This work is supported by the Instituto de Salud Carlos III, Government of Spain and FEDER funds of the European Union through the DTS18/00136 research projects and by the Ministerio de Ciencia, Innovación y Universidades, Government of Spain through the RTI2018-095894-B-I00 research projects, as well as through Ayudas para la formación de profesorado universitario (FPU), Ref. FPU18/02271. Also, this work has received financial support from the European Union (European Regional Development Fund - ERDF) and the Xunta de Galicia, Centro de Investigación del Sistema Universitario de Galicia, Ref. ED431G 2019/01.

